# High levels of all-cause mortality among people who inject drugs in Greece in 2018–2022

**DOI:** 10.1101/2022.10.07.22280788

**Authors:** Sotirios Rousssos, Theodoros Angelopoulos, Evangelos Cholongitas, Spyridon Savvanis, Nikolaos Papadopoulos, Andreas Kapatais, Athina Hounta, Panagiota Ioannidou, Melani Deutsch, Spilios Manolakopoulos, Vasileios Sevastianos, Maria-Vasiliki Papageorgiou, Ioannis Vlachogiannakos, Maria Mela, Ioannis Elefsiniotis, Spyridon Vrakas, Dimitrios Karagiannakis, Fani Pliarchopoulou, Savvas Chaikalis, Effrosyni Tsirogianni, Mina Psichogiou, Georgios Kalamitsis, Foteini Leobilla, Dimitrios Paraskevis, Meni Malliori, Ioannis Goulis, Georgios Papatheodoridis, Angelos Hatzakis, Vana Sypsa

## Abstract

**Aims:** To estimate all-cause mortality in a cohort of people who inject drugs (PWID) over the period 2018–2022 in the two major cities of Greece (Athens and Thessaloniki).

**Methods:** PWID were recruited from two community-based seek-test-treat programs for HCV and HIV infections in Athens and Thessaloniki. Participants provided information on sociodemographic characteristics, injection practices, social security number, access to harm reduction and treatment. Data on the vital status and date of death were identified from the national HCV treatment registry. All-cause mortality rates (CMR) were estimated as well as age-, gender- and calendar-year standardized mortality ratios (SMR). Determinants of mortality were assessed using a Cox proportional-hazards model.

**Results:** Of 2,433 PWID, 243 (10.0%) died over a total of 6,649 person-years (PYs) of follow-up. The CMR [95% confidence interval (CI)] was 3.50 (3.08–3.97) deaths per 100 PYs; 3.03 (2.58–3.57) in Athens and 4.56 (3.74–5.57) in Thessaloniki. An increasing trend in CMR was identified over the period 2018–2022 in Athens (p for trend=0.004). The overall SMR (95% CI) was 17.17 (15.14–19.47) per 100 PYs in the combined cohort; 15.10 (12.85–17.75) in Athens and 21.72 (17.78–26.53) in Thessaloniki. The SMR was particularly increased in younger ages, females, those injecting daily, and HIV-infected PWID. Older age, living in Thessaloniki, Greek origin, homelessness, daily injecting drug use, HIV, and HCV infections were independently associated with all-cause mortality.

**Conclusion:** All-cause mortality among PWID in Greece during 2018–2022 is high with the population in Thessaloniki being particularly affected. The increasing trend in mortality in Athens might be the long-term impact of the pandemic on the health of PWID. Preventive programs such as take-home naloxone distribution and community involvement to increase harm reduction, screening, and uptake of antiretroviral and chronic hepatitis C treatment are urgently needed.

## INTRODUCTION

People who inject drugs (PWID) represent a socially disadvantaged and vulnerable population with excess risk of morbidity and mortality as it has been demonstrated in many studies and metanalyses (1, 2). The all-cause crude mortality rate among people using extramedical opioids ranges from 0.80 to 1.61 per 100 person-years (PYs) in high-income regions based on the Global Burden of Disease regional classification system (Australasia, Central and Western Europe, and North America) (2). The excess mortality varies and is 6–16 times higher relative to the age- and sex-matched general population. The major causes of mortality are poisoning by drugs, infectious diseases, including HIV and HCV infection, suicide and unintentional injuries, liver, cancer, cardiovascular, respiratory diseases, etc. Fatal drug overdose is the major cause of death in high-income countries (2) and is the dominant cause of death among individuals aged 25–44 years in the US (3). HIV infection used to be the major cause of death (4, 5) although, after the widespread use of combination antiretroviral therapy (cART), the proportion of deaths attributable to HIV was reduced while drug poisoning emerged as the main cause of death (6, 7).

In Athens, Greece, a major HIV outbreak among PWID occurred between 2011–2013, which is considered the largest epidemic in the 21st century among PWID in Western Europe and North America (8, 9). The outbreak was controlled by the implementation of a large seek–test–treat program (ARISTOTLE) and enhanced provision of harm reduction services (needle and syringe programs, opioid substitution treatment). Indeed, HIV incidence declined from the peak of 7.8/100 PYs in 2012 to 1.7/100 PYs by the end of 2013 and remained stable until 2020 (10, 11). Not surprisingly, due to the ongoing transmission, the prevalence increased from 14% in 2013 to 22% in 2020 in a cohort of repeatedly tested PWID in these two periods, as it was demonstrated in a second similar program (ARISTOTLE HCV-HIV) implemented during 2018–2020 (11). Recently, an HIV outbreak occurred in Thessaloniki (the second-largest city in Greece) during 2019–2021, based on the data collected from another community-based intervention (ALEXANDROS program) (12).

These two recent HIV epidemics among PWID in Greece were the motive to investigate mortality in this population using data from the two interventions implemented in Athens and Thessaloniki. The aim of this study is to provide an estimate of the all-cause mortality among PWID in Greece during the period 2018–2022, to compare it to that of the general population as well as to estimates from PWID populations in other high-income countries, and to identify determinants of mortality.

## METHODS

### Design of ARISTOTLE HCV-HIV and ALEXANDROS programs

ARISTOTLE HCV-HIV (2018–2020) and ARISTOTLE (2019–2021) were community-based programs aimed at increasing the diagnosis and linkage to care for HIV and/or HCV in Athens and Thessaloniki, respectively. The two programs had similar design and eligibility criteria. Participants should have been older than 18 years with a history of injecting drug use and resided in the city where each program was implemented.

Respondent-driven sampling (RDS) was used to recruit participants. RDS is a peer-driven chain referral where recruitment begins with a limited number of initial recruits (‘seeds’); individuals receive coupons and are asked to draw from their existing injection networks to identify up to three to five potential recruits, who then present themselves to the program site. A dual-monetary incentive system was used, in which participants received incentives for participating as well as for recruiting others. The programs were conducted in multiple recruitment rounds. Participants could participate in all recruitment rounds, but only once per round. After obtaining written informed consent, personal interviewing was used to collect information on participants’ socio-demographic characteristics, injection, and sexual behaviour, social security number as well as on access to HIV/HCV testing, treatment, and prevention programs. Participants were tested for HIV and HCV and patients were linked to care. Linkage to HCV care and treatment was possible through the provision of the social security number as this is a prerequisite for patient enrollment to the National Hepatitis C Treatment Registry and for accessing direct acting antivirals for free. Study personnel helped PWID without social security number to issue one.

### Outcome measures

The main outcome measure was all-cause mortality over the period April 2018 – June 2022. The crude mortality rate (CMR) in each cohort as well as the combined mortality rate from both cohorts were presented as deaths per 100 PYs. The program collected information on participants’ vital status and date of death from the National Hepatitis C Treatment Registry using their social security numbers. The follow-up time was calculated as the time between the date of the first participation in the program and the date of death, or the date of last known vital status (June 12, 2022).

The second outcome measure was the Standardized Mortality Ratio (SMR). This index allows to compare the mortality that would be expected in the PWID population given national death rates for the adult population with similar age and gender. SMRs with 95% confidence intervals (CIs) were estimated for the total person-years of follow-up using indirect standardisation, as the number of observed deaths divided by the expected number of deaths in the general population, according to age-, sex-, and calendar year-specific death rates in Greece obtained from The Human Mortality Database (13). As there were no data after 2019, we have used the 2019 mortality rates for the years 2020–2022.

### Statistical analysis

Categorical variables were described using frequencies and percentages. Continuous variables were described using the appropriate measures of central tendency and spread [means and standard deviation (SD) or median, 25th, and 75th percentiles]. In order to compare participants’ baseline characteristics (as assessed in their first visit), the chi-square test, t-test, or Mann–Whitney U-test were used, as appropriate.

Trends in all-cause mortality rates over calendar year were obtained from a Poisson regression model. Univariable and multivariable Cox proportional hazard models were used to assess determinants of mortality and the associated hazard ratios (HR) and 95% 95% CIs were obtained. Graphical methods as well as tests based on the Schoenfeld residuals were used to check the assumption of proportionality of the hazards. Collett’s approach was applied for model selection and likelihood ratio tests were used for variable inclusion/exclusion decisions. In the final model, age was centred at the mean value (40 years).

As the information on vital status could be retrieved only for PWID with an available social security number, to assess for selection bias, we compared the characteristics of PWID who had a social security number with those who had not.

Ten PWID participated in both programs. They were included only once in the analysis using the information of their first participation. Analyses were conducted using Stata version 17.0.

### Ethics

ARISTOTLE HCV-HIV program was approved by the Institutional Review Board of Athens University Medical School and of the Hellenic Scientific Society for the Study of AIDS, STDs, and Emerging Diseases. ALEXANDROS program was approved by the Institutional Review Board of Athens University Medical School and of the Hellenic Scientific Society for the Study of AIDS, STDs, and Emerging Diseases. Persons who were eligible to participate based on the eligibility screening process were asked to provide written informed consent. The informed consent form included information about the program, explained that confidentiality would be protected and that participants were free to withdraw their consent at any point of the process. The questionnaire and the blood sample were linked through the RDS coupon number. The name of the participant was not recorded in the questionnaire database or on the blood collection tube. The correspondence of the name and coupon number was recorded in a separate file that could only be accessed by authorized members of the research team.

## RESULTS

### Participants

ARISTOTLE HCV-HIV recruited 1,634 PWID during April 2018–March 2020. ALEXANDROS recruited 1,101 participants during September 2019–July 2021 (2,725 unique participants after keeping only the first record of ten PWID who participated in both programs). Data on social security number was missing in 292 out of 2,725 unique participants (10.7%). Thus, information on the last known vital status was available for 2,433 PWID; 1,386/1,634 (84.8%) and 1,047/1,091 (96.0%) participants of ARISTOTLE HCV-HIV and ALEXANDROS, respectively.

In the combined cohort (n = 2,433), the mean age (SD) was 40.1 (8.3) years, 84.6% were males, 89.2% were of Greek origin, and 37.2% had completed high school or higher education (Table 1). Current homelessness and history of imprisonment in the past 12 months were reported by 19.1% and 10.7%, respectively. One out of five participants (22.2%) reported extensive drug injection networks (≥30 people). Concerning injection behaviour in the past 12 months, 70.3% reported heroin/thai as the main substance used, 27.4% injected daily and 20.8% reported receptive syringe sharing. The majority of participants (65.0%) were active injectors (injection in the past 30 days). Current use of opioid substitution treatment programs was reported by 24.0%. HIV and anti-HCV prevalence were 12.4% and 72.8%, respectively, and 11.8% were HIV/HCV-coinfected (Table 1, Supplementary Table 1).

**Table 1.** Crude mortality rate (CMR) and standardized mortality ratios (SMR) by socio-demographic and network characteristics, drug use behaviour, access to opioid substitution treatment, and HIV/HCV status in people who inject drugs (as assessed in the first visit to the programs).

| **Variables** | **n (%)** | **No. Deaths (%)** | **PYs of Follow-up** | **CMR (/100 PYs) (95% CI)** | **SMR (95% CI)** |
| --- | --- | --- | --- | --- | --- |
| **Overall** | **2433 (100.0)** | **243 (10.0)** | **6949.4** | **3.50 (3.08–3.97)** | **17.17 (15.14–19.47)** |
| City |  |  |  |  |  |
| Athens | 1386 (57.0) | 147 (10.6) | 4845.6 | 3.03 (2.58–3.57) | 15.10 (12.85–17.75) |
| Thessaloniki | 1047 (43.0) | 96 (9.2) | 2103.8 | 4.56 (3.74–5.57) | 21.72 (17.78–26.53) |
| Age (years) |  |  |  |  |  |
| [18–35) | 615 (25.3) | 44 (7.2) | 1767.0 | 2.49 (1.85–3.35) | 35.64 (26.53–47.90) |
| [35–50) | 1520 (62.5) | 148 (9.7) | 4345.9 | 3.41 (2.90–4.00) | 20.58 (17.52–24.18) |
| [50–71) | 298 (12.2) | 51 (17.1) | 836.6 | 6.10 (4.63–8.02) | 8.91 (6.77–11.72) |
| Gender |  |  |  |  |  |
| Female | 375 (15.4) | 27 (7.2) | 1118.3 | 2.41 (1.66–3.52) | 28.28 (19.39–41.24) |
| Male | 2057 (84.6) | 216 (10.5) | 5829.9 | 3.71 (3.24–4.23) | 16.37 (14.32–18.70) |
| Country of origin |  |  |  |  |  |
| Greece | 2167 (89.2) | 229 (10.6) | 6260.2 | 3.66 (3.21–4.16) | 17.95 (15.77–20.43) |
| Other | 263 (10.8) | 13 (4.9) | 684.2 | 1.90 (1.10–3.27) | 9.36 (5.44–16.12) |
| Highest level of education completed |  |  |  |  |  |
| High school or higher | 902 (37.2) | 82 (9.1) | 2707.9 | 3.03 (2.44–3.76) | 16.31 (13.13–20.25) |
| Middle/secondary school or below | 1520 (62.8) | 159 (10.5) | 4220.0 | 3.77 (3.23–4.40) | 17.50 (14.98–20.44) |
| Currently homeless |  |  |  |  |  |
| No | 1965 (80.9) | 176 (9.0) | 5558.0 | 3.17 (2.73–3.67) | 16.11 (13.90–18.68) |
| Yes | 463 (19.1) | 66 (14.3) | 1382.0 | 4.78 (3.75–6.08) | 20.57 (16.16–26.19) |
| Currently health insurance |  |  |  |  |  |
| Yes | 483 (20.0) | 48 (9.9) | 1494.7 | 3.21 (2.42–4.26) | 15.32 (11.54–20.32) |
| No | 1929 (80.0) | 194 (10.1) | 5399.3 | 3.59 (3.12–4.14) | 17.71 (15.39–20.39) |
| Unemployed |  |  |  |  |  |
| Yes | 2130 (87.8) | 212 (10.0) | 6161.1 | 3.44 (3.01–3.94) | 16.93 (14.80–19.37) |
| No | 296 (12.2) | 30 (10.1) | 773.5 | 3.88 (2.71–5.55) | 18.70 (13.08–26.75) |
| Imprisonment (past 12 months) |  |  |  |  |  |
| No | 2168 (89.3) | 211 (9.7) | 6170.6 | 3.42 (2.99–3.91) | 16.72 (14.61–19.14) |
| Yes | 260 (10.7) | 31 (11.9) | 769.5 | 4.03 (2.83–5.73) | 20.49 (14.41–29.14) |
| Size of participant's network |  |  |  |  |  |
| ≤10 | 943 (39.1) | 88 (9.3) | 2524.0 | 3.49 (2.83–4.30) | 16.98 (13.78–20.93) |
| 11–30 | 932 (38.7) | 91 (9.8) | 2630.2 | 3.46 (2.82–4.25) | 17.46 (14.22–21.44) |
| 30+ | 534 (22.2) | 63 (11.8) | 1724.1 | 3.65 (2.85–4.68) | 17.63 (13.77–22.56) |
| Main substance of use |  |  |  |  |  |
| Heroin/Thai | 1705 (70.3) | 176 (10.3) | 4613.6 | 3.81 (3.29–4.42) | 18.12 (15.64–21.01) |
| Cocaine/Speedball | 675 (27.8) | 65 (9.6) | 2196.6 | 2.96 (2.32–3.77) | 15.38 (12.06–19.62) |
| Other | 45 (1.9) | 1 (2.2) | 125.3 | 0.8 (0.11–5.66) | 5.29 (0.75–37.56) |
| Injecting drug use (past 30 days) |  |  |  |  |  |
| No | 850 (35.0) | 65 (7.6) | 2165.7 | 3.00 (2.35–3.83) | 13.56 (10.63–17.29) |
| Yes | 1578 (65.0) | 177 (11.2) | 4774.3 | 3.71 (3.20–4.30) | 18.96 (16.36–21.96) |
| Injecting drug use (past 12 months) |  |  |  |  |  |
| No | 344 (14.2) | 19 (5.5) | 768.2 | 2.47 (1.58–3.88) | 9.35 (5.97–14.66) |
| Yes | 2084 (85.8) | 223 (10.7) | 6171.9 | 3.61 (3.17–4.12) | 18.43 (16.16–21.02) |
| Duration of injecting drug use (years) |  |  |  |  |  |
| <18 | 1111 (46.0) | 97 (8.7) | 3219.0 | 3.01 (2.47–3.68) | 24.60 (20.16–30.02) |
| ≥18 | 1305 (54.0) | 145 (11.1) | 3683.4 | 3.94 (3.35–4.63) | 14.30 (12.16–16.83) |
| Daily injecting drug use |  |  |  |  |  |
| No | 1759 (72.6) | 146 (8.3) | 5055.4 | 2.89 (2.46–3.40) | 13.51 (11.49–15.89) |
| Yes | 663 (27.4) | 95 (14.3) | 1870.1 | 5.08 (4.15–6.21) | 28.98 (23.70–35.43) |
| Receptive syringe sharing |  |  |  |  |  |
| Never | 1914 (79.2) | 180 (9.4) | 5385.3 | 3.34 (2.89–3.87) | 15.80 (13.66–18.29) |
| At least once | 502 (20.8) | 61 (12.2) | 1509.5 | 4.04 (3.14–5.19) | 23.24 (18.08–29.86) |
| Use drugs divided with a syringe that someone else had already injected with |  |  |  |  |  |
| Never | 2006 (82.8) | 183 (9.1) | 5702.7 | 3.21 (2.78–3.71) | 15.47 (13.38–17.88) |
| At least once | 416 (17.2) | 59 (14.2) | 1216.9 | 4.85 (3.76–6.26) | 26.16 (20.27–33.76) |
| Currently on OST |  |  |  |  |  |
| Yes | 583 (24.0) | 61 (10.5) | 1746.1 | 3.49 (2.72–4.49) | 12.57 (9.78–16.15) |
| No | 1842 (76.0) | 181 (9.8) | 5184.9 | 3.49 (3.02–4.04) | 19.53 (16.89–22.60) |
| Anti-HIV |  |  |  |  |  |
| (–) | 2130 (87.6) | 190 (8.9) | 6017.2 | 3.16 (2.74–3.64) | 15.11 (13.10–17.42) |
| (+) | 301 (12.4) | 53 (17.6) | 925.6 | 5.73 (4.37–7.50) | 34.12 (26.07–44.66) |
| Anti-HCV |  |  |  |  |  |
| (–) | 661 (27.2) | 42 (6.4) | 1694.9 | 2.48 (1.83–3.35) | 12.20 (9.01–16.50) |
| (+) | 1770 (72.8) | 201 (11.4) | 5247.9 | 3.83 (3.34–4.40) | 18.81 (16.38–21.60) |
| First participation<br>based on the first<br>national lockdown<br>(15/3/2020) |  |  |  |  |  |
| Before | 1953 (80.3) | 212 (10.9) | 6213.7 | 3.41 (2.98–3.90) | 16.60 (14.51–19.00) |
| After | 480 (19.7) | 31 (6.5) | 735.8 | 4.21 (2.96–5.99) | 22.38 (15.74–31.82) |
| Year of entry (first<br>participation) to the<br>study |  |  |  |  |  |
| 2018 | 1086 (44.6) | 124 (11.4) | 4070.4 | 3.05 (2.55–3.63) | 15.35 (12.88–18.31) |
| 2019 | 682 (28.0) | 74 (10.9) | 1731.9 | 4.27 (3.40–5.37) | 19.66 (15.65–24.69) |
| 2020 | 508 (20.9) | 34 (6.7) | 976.1 | 3.48 (2.49–4.88) | 16.70 (11.93–23.37) |
| 2021 | 157 (6.5) | 11 (7.0) | 171.1 | 6.43 (3.56–11.61) | 39.85 (22.07–71.95) |
| Abbreviations: OST-Opioid substitution treatment |  |  |  |  |  |

There were differences in the characteristics of the participants of the two programs. A higher proportion of ARISTOTLE HCV-HIV participants in Athens were females, with a higher educational level, homeless, unemployed, active injectors, and enrolled in OST programs (Supplementary Table 1). In addition, they reported a higher frequency of imprisonment and larger networks of drug use. A lower proportion of migrants participated in ARISTOTLE HCV-HIV as compared to ALEXANDROS. HIV prevalence was 17.1% in Athens and 6.2% in Thessaloniki, and anti-HCV prevalence was 79.9% and 63.4%, respectively (Supplementary Table 1).

In Supplementary Table 2, we compare the characteristics of PWID included in the analysis with those who were not, due to lack of data on their social security number. PWID without this information were more often younger, of non-Greek origin, unemployed, active users, with a lower level of education, unstable accommodation, reporting more often a history of imprisonment in the past 12 months, daily injection, higher network size, and sharing syringes, as compared to PWID with available social security number.

### Crude Mortality Rates

Participants contributed a total of 6,649 PYs of follow-up. Over the study period (April 2018 – June 2022), 243 (10.0%) deaths were recorded in 2,433 PWID in both cities. The CMR (95% CI) was 3.50 (3.08–3.97) deaths per 100 Pys. Higher CMRs were identified in participants from Thessaloniki, aged ≥50 years, currently homeless, with daily injecting drug use, who used drugs divided with a syringe that someone else had already injected with as well as PWID with HIV infection and HIV-HCV coinfection (Table 1).

When analyzing each city separately, 147 and 96 deaths were recorded in Athens and Thessaloniki over a total follow-up of 4,846 Pys and 2,104 Pys, respectively. The CMR (95% CI) was 3.03 (2.58–3.57) in Athens and 4.56 (3.74–5.57) in Thessaloniki (Table 1).

An increasing trend in overall mortality rates over calendar year was identified (p-value for trend <0.001) (Figure 1, Supplementary Table 3, Supplementary Figure 1). This finding reflects the increasing trend in mortality among PWID in Athens over the period 2018–2022 (p-value for trend=0.004). The CMR in Athens in the period January-June 2022 was statistically significantly higher than that estimated in 2019 or in 2018-2019 combined. No statistically significant trend over calendar year was identified in Thessaloniki (p-value for trend=0.475).

**Figure 1.**
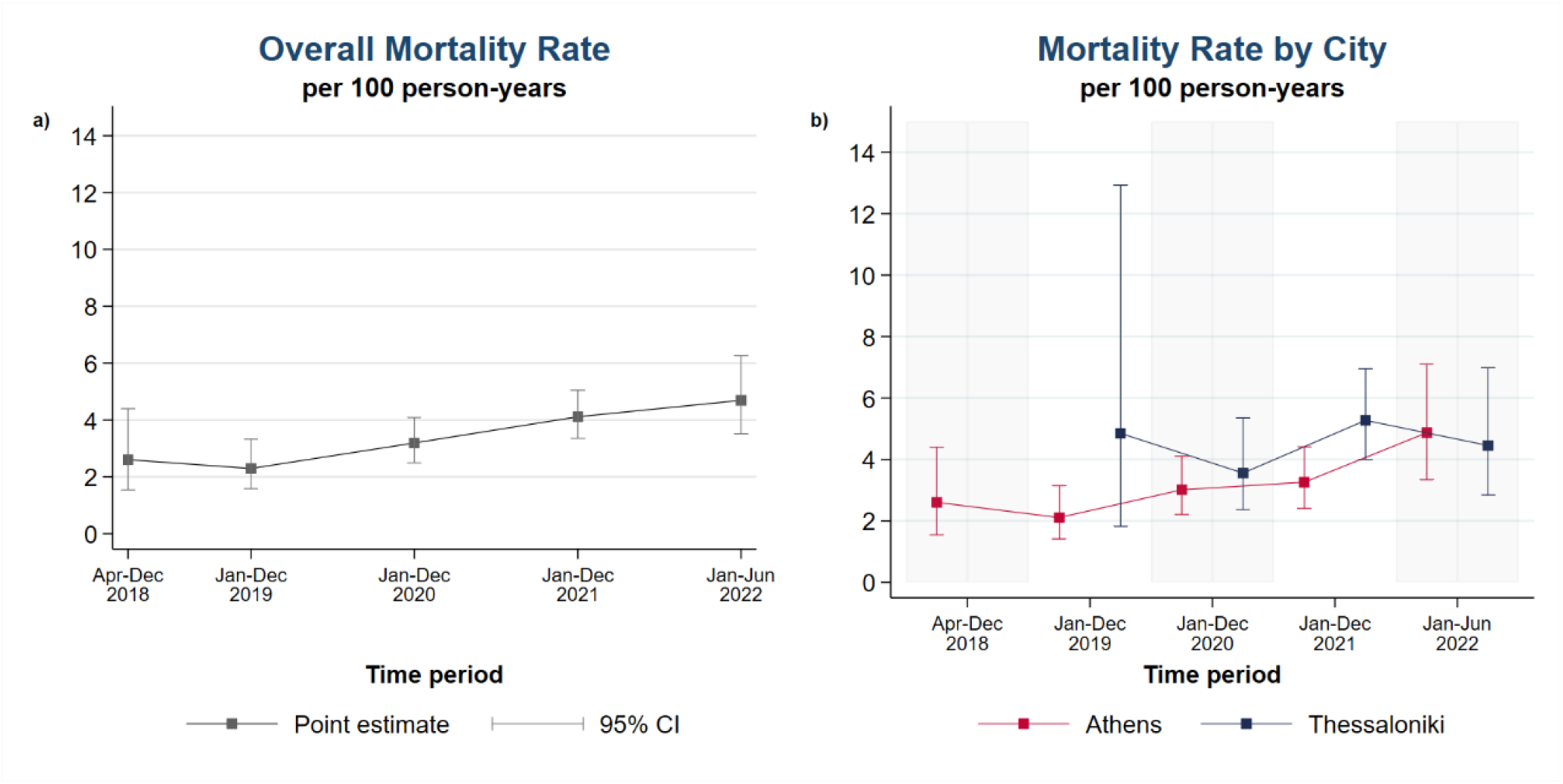
All-cause mortality rate (/100 person-years) among people who inject drugs in Greece over the period 2018–2022 by calendar year (a) overall, (b) by city (Athens, Thessaloniki)

### Standardized Mortality Ratios

The overall age-, gender- and calendar-year SMR (95% CI) was 17.17 (15.14–19.47) per 100 PYs in the combined cohort (Table 1). The SMR (95% CI) in Athens and Thessaloniki was 15.10 (12.85–17.75) and 21.72 (17.78–26.53), respectively.

The SMR was higher than one – i.e. mortality was higher than that of the general population - in all subgroups of the PWID population. It increased dramatically in younger ages to 35.64 (95% CI: 26.53–47.90) and 20.58 (95% CI: 17.52–24.18) for PWID aged 18–35 and 35–49 years, respectively [8.91 (95% CI: 6.77–11.72) for PWID aged 50–71 years]. Female PWID had a higher SMR than male (28.28 vs. 16.37). The SMR for PWID injecting daily was 28.98 (95% CI: 23.70–35.43) versus 13.51 (95% CI: 11.49–15.89) for those injecting less frequently. HIV-infected PWID had more than double SMR as compared to seronegative PWID [34.12 (95% CI: 26.07–44.66) vs. 12.90 (95% CI: 9.01–16.50)] (Table 1).

### Risk factors for mortality in PWID

Univariable and multivariable analysis of risk factors for all-cause mortality are presented in Table 2. Sociodemographic factors including older age, Greek origin, lack of stable accommodation, as well as daily injection, HIV and HCV infections were identified as independent risk factors for death.

**Table 2.**
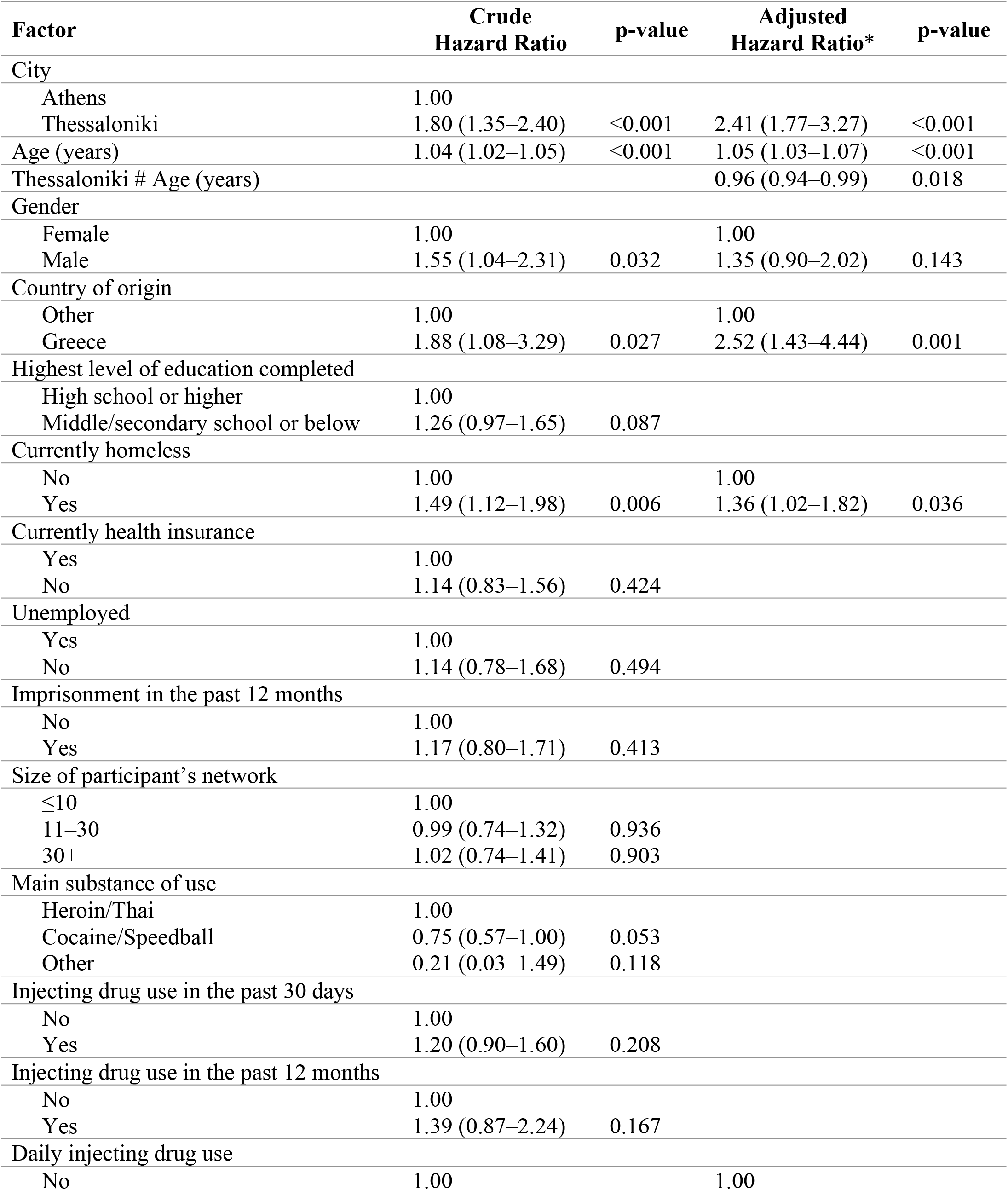
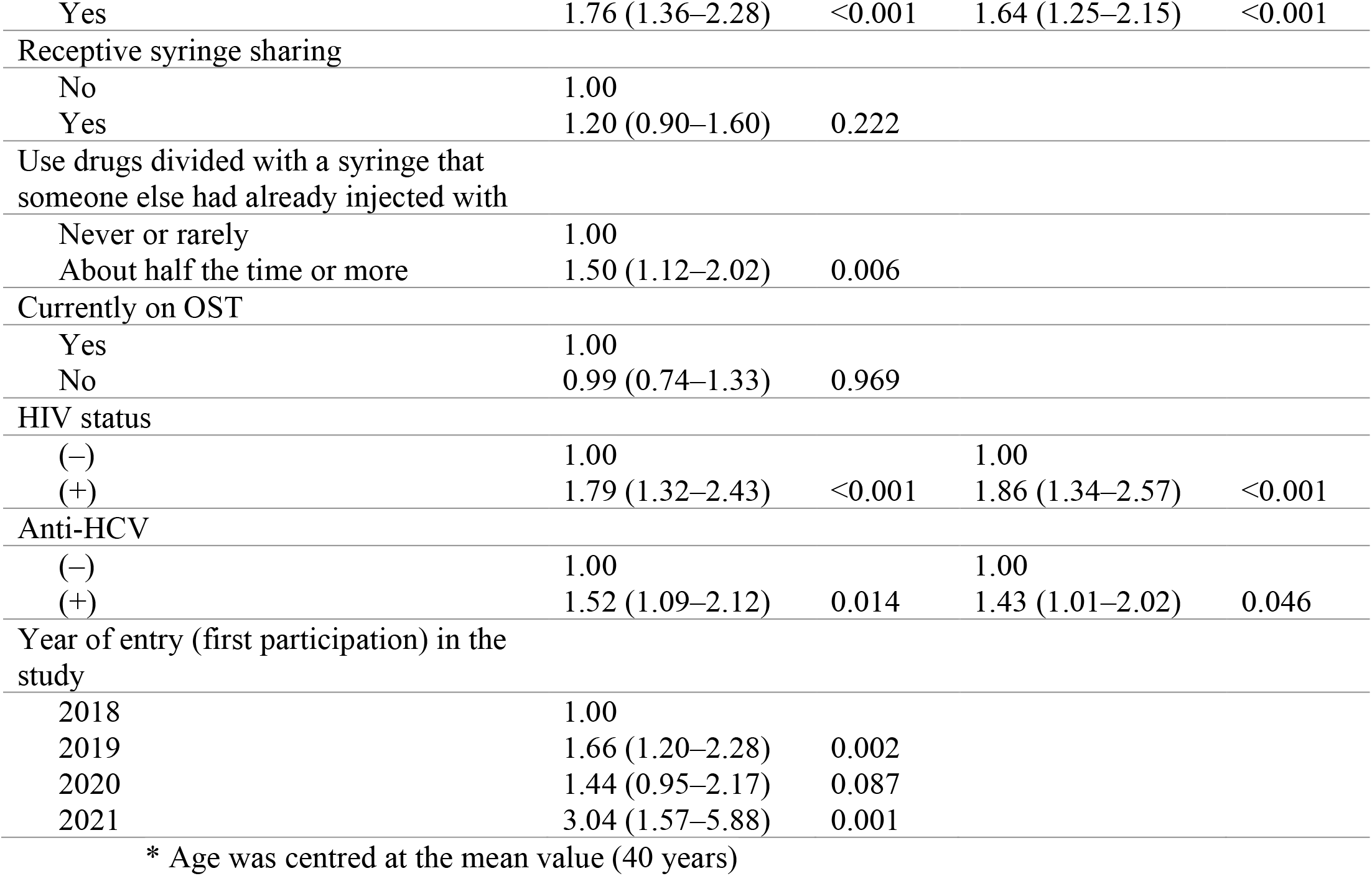
Determinants of all-cause mortality among people who inject drugs (Cox proportional hazards models, univariable and multivariable analysis).

In the adjusted analysis, PWID living in Thessaloniki had 2.4 times higher risk of death as compared to those living in Athens [2.41 (95% CI: 1.77–3.27, p<0.001)] (for PWID at the age of 40 years). An additional year of age increased the hazard of dying by 5%. A significant interaction between city and age was identified; the gap in the risk of death between Thessaloniki and Athens decreased with increasing age. Other variables associated with higher hazard were Greek origin (vs. other: HR [95% CI]: 2.52 [1.43– 4.44]) and homelessness (vs. not homeless: HR [95% CI]: 1.36 [1.02–1.82]). Concerning injection practices, PWID reporting daily injecting in the past 12 months had 1.64 times higher risk of death as compared to those injecting less frequently (95% CI: 1.25–2.15, p<0.001). Furthermore, HIV-infected PWID, as well as PWID with exposure to HCV had 1.86- and 1.43-times higher risk of death as compared to those without HIV and HCV, respectively (Table 2).

## DISCUSSION

In this study, we document high levels of all-cause mortality in PWID in Greece over the period 2018–2022. Our sample comprised community-recruited people who injected drugs with two-thirds of them reporting injection in the past 30 days. The all-cause crude mortality (95% CI) was 3.50 (3.08–3.97) per 100 PYs, and it was considerably higher in Thessaloniki as compared to Athens (4.56 vs. 3.03, respectively, with non-overlapping 95% CIs). After adjusting for age and gender, mortality within the cohort was 17 times higher than in the general Greek population.

The magnitude of all-cause mortality documented in this study is the highest observed in Western and Central Europe, North America, Australia, and East Asia based on a recent metanalysis of 99 studies conducted in the last 40 years among people using extramedical opioids (2). Specifically, the all-cause crude mortality of 3.50 (3.08–3.97) per 100 PYs estimated in our analysis was higher compared with 1.56 (1.32–1.84) in Western Europe (54 studies), 1.17 (0.61–2.23) in Central Europe (3 studies), 1.61 (1.36–1.91) in North America (20 studies), 0.80 (0.67–0.96) in Australasia (7 studies), and 1.80 (1.31–2.35) in East Asia (6 studies) (Figure 2). Similar levels of mortality with Greece were noted in North Africa and the Middle East [3.40 (0.97–11.87) (2 studies)] and Southeast Asia [4.53 (3.11–6.62) (3 studies)]. Only two studies in South Asia report a higher estimate [7.62 (4.84–12.00) based on 2 studies from Bangladesh] (Figure 2). In addition, the mortality rate among PWID in Greece was higher than the corresponding estimate when only studies in cohorts of people who inject opioids were considered [2.71 (2.14–3.42)] but with overlapping 95% Cis (2). It should be noted that the widespread uptake of cART has resulted in a decline in HIV deaths among PWID after 1997, as in e.g. Baltimore, where a dramatic drop has been observed (7). As a result, the comparison of mortality rates in various settings should take this into account. A more thorough assessment of the individual studies in cohorts of people who inject opioids included in the metanalysis reveals that all those reporting higher mortality estimates than ours are either from Asia (5 studies) or from Europe, but derived from a high-risk population of PWID attending an emergency department (1 study) or based on data collected prior to the introduction of cART (1 study) (2).

**Figure 2.**
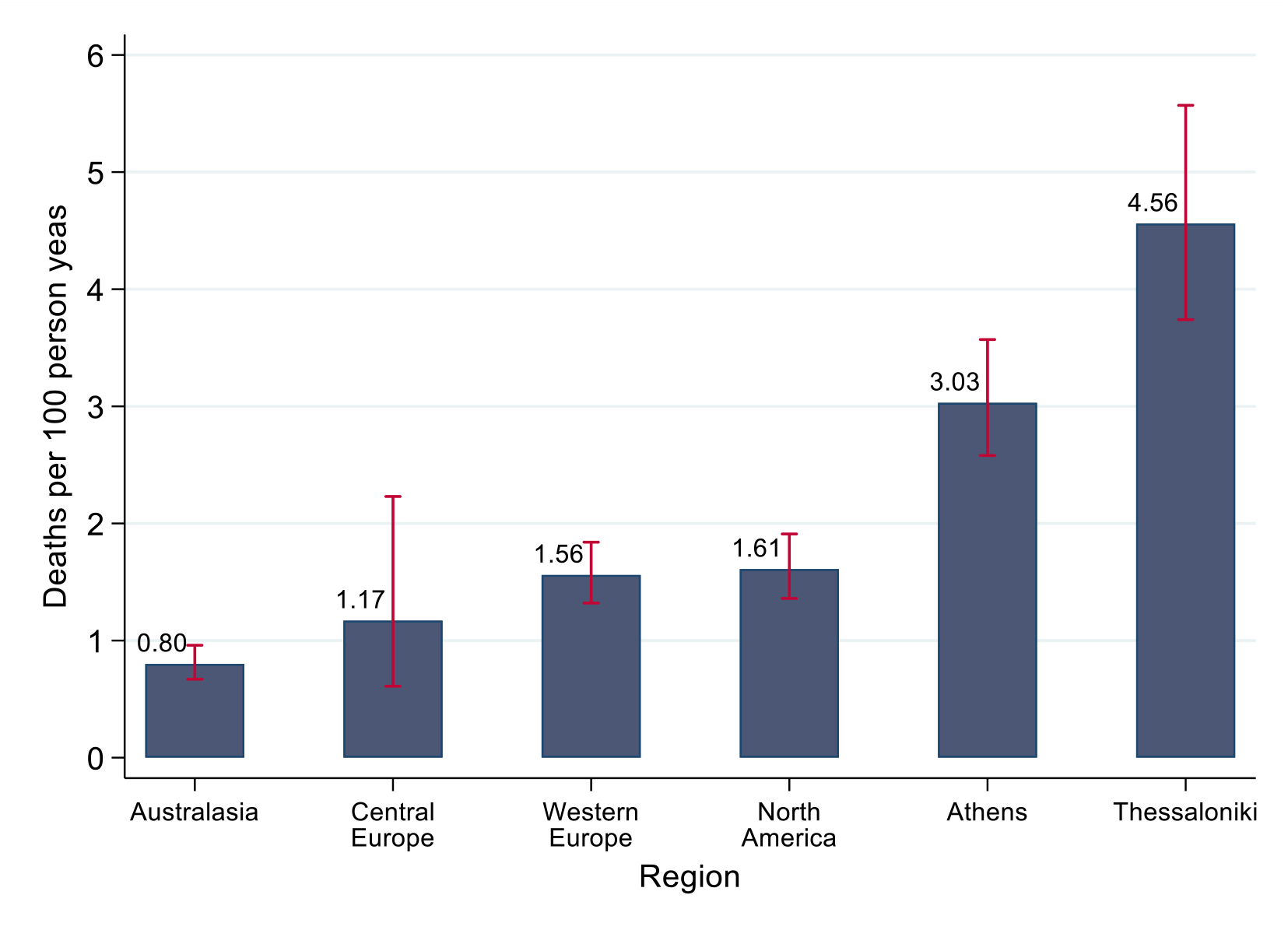
All-cause mortality (/100 person-years) among people who inject drugs in Athens and Thessaloniki, Greece, over the period 2018–2022 compared with the corresponding estimates by region from a metaanalysis of studies in people using extramedical opioids in developed countries (red lines: 95% Confidence Interval) (2)

The overall SMR (95% CI) of 17.17 (15.14–19.47) reported in our study that compares the mortality among PWID to that of the age- and sex-matched general population was also higher than that reported in the metanalysis [10.03 (7.64– 13.17)] and at similar levels when ten studies specifically for people who inject opioids were included [16.37 (10.92–24.55)]. A more detailed analysis of the SMR in the various subgroups of our cohort revealed that the youngest age group in the study (18–35 years old) was over 35 times more likely to die compared to their age- and sex-matched general population in Greece. Similarly, female PWID, PWID living in Thessaloniki, those injecting daily and HIV infected had particularly high SMR. Women and younger individuals had a higher SMR in the meta-analysis of studies among people using extramedical opioids, but the 95% confidence intervals overlapped. There was a large gap in the SMRs in HIV-positive and HIV-negative PWID in our analysis (34.12 vs. 15.11). Although a study among PWID recruited between 2014–2016 in Vietnam (14) identified a similar gap in the age- and gender-SMR among HIV-negative and HIV-positive PWID [4.2 (3.0–5.8) and 12.0 (9.3–15.1), respectively], the mortality gap between PWID and the general population was much more narrow than that identified in Greece.

The causes of high mortality among PWID in Greece are not known. Based on the analysis of risk factors, older age, living in Thessaloniki, Greek origin, homelessness, daily injecting drug use, HIV and HCV infections were independently associated with all-cause mortality. The recent HIV epidemics in Athens and Thessaloniki (11, 15) suggest that HIV infection may have a prominent role in the increased mortality. The role of drug poisoning and opioid overdose cannot be documented without data from a study on cause-specific mortality. Based on the National Documentation and Information Centre for Drugs and Drug Addiction, 242 opioid overdose deaths were reported in 2018 (16, 17) suggesting a prominent role of overdose deaths in PWID mortality. The finding of 50% higher mortality in Thessaloniki, as compared to Athens, is striking, in particular considering that the profile of the population was more favourable (the prevalence of factors associated with increased risk of death – such as homelessness, HIV infection, non-Greek origin – was lower in Thessaloniki). The gap in the risk of death between Thessaloniki and Athens was more apparent in younger ages. It should be noted that the harm reduction programs in Greece mainly focus on the Athens population due to the HIV outbreak recorded in 2011. Based on participants’ self-reports, the proportion of PWID currently on OST and of those having received syringes is lower in Thessaloniki than in Athens. Ninety-three percent of syringes distributed in 2019 were provided in Athens (18). Furthermore, the number of distributed syringes in 2020 in Greece was reduced by 17%, compared to 2019, because of the suspension of these programs due to the COVID-19 pandemic (19). There may be additional reasons for the higher mortality in Thessaloniki that should be explored, such as overdose and type/quality of drugs circulating in this population.

Several studies have shown that retention in opioid substitution treatment is associated with substantial reductions in the risk for all cause mortality in people dependent on opioids (20, 21). However, we did not identify such an association in our study. A possible explanation might be that participants on OST who were reached by these community-based programs with peer-driven chain referral might have been high-risk individuals. Indeed, the majority of participants on OST (62.2%) reported injecting drug use in the past 30 days.

We identified an increasing trend in all-cause mortality among PWID over the period 2018–2022. This finding is attributed to an increase only in Athens where a significantly higher CMR was identified in 2022 as compared to 2018-2019. We cannot rule out the possibility that the COVID-19 pandemic may have contributed to this increase. We did not observe a sharp increase in 2020 as in e.g. the US where the annual incidence of fatal overdose increased by 30% between 2019 and 2020 (22). However, we may be seeing the longer-term impact of the pandemic on the health and wellbeing of PWID as it had been hypothesized early on (23-25). The health consequences of “big events”, such as the COVID-19 pandemic, have been found to disproportionately affect poor and disadvantaged populations, including those using drugs (26, 27). Athens is the largest metropolitan centre in Greece, with a high population density and PWID have a higher prevalence of homelessness and unemployment as compared to Thessaloniki.

Our study has some limitations. Information on mortality was available for PWID with available social security number. PWID without this information could not be included; they were more often younger, of non-Greek origin, unemployed, active users, with lower level of education, unstable accommodation, reporting more often history of imprisonment in the past 12 months, daily injection, higher network size, sharing syringes as compared to PWID included in the analysis. As a result, the overall mortality rate and SMR may be underestimated. However, considerable efforts were made by the personnel of the program to help PWID to identify their social security number (in case they did not remember it/did not know they had one), or even to issue one. As a result, only 10.7% of the original sample was excluded due to lack of this information. Other limitations of the study are the relatively short follow-up and the lack of data on cause-specific mortality. The latter could provide a more complete picture of the drivers affecting mortality among PWID in Greece. However, national databases on mortality are slow to include updated cause-specific data in most countries and Greece. Finally, it should be pointed out that most of the study period coincided with the COVID-19 pandemic where access to hospitals and medical services was limited. Data from the US indicate an increase in opioid overdose deaths during the pandemic (22, 28, 29).

This study has some strengths such as the timeliness of mortality data, a large sample size and well-established enrollment frame able to recruit PWIDs with high coverage (10). Based on the 2020 official population size estimation of current injectors (n = 2,488) in Greece (19) and the corresponding number of current injectors included in our analysis (n = 1,578), the study sample represents 63.42% of the high-risk PWID population. To our knowledge, this is the first study reporting mortality in PWID in Greece.

The high mortality of PWID in Greece, in particular in Thessaloniki, and the increasing trends in Athens are alarming and a call to action for health authorities, the scientific and PWID communities. Preventive programs such as take-home naloxone distribution as overdose is one of the most frequent causes of death among PWID (2, 7, 30) - and community involvement to increase harm reduction, screening and uptake of antiretroviral and chronic hepatitis C treatment are urgently needed.

## Data Availability

Data are not available for sharing.

---

**Supplementary Table 1.** Socio-economic and network characteristics, drug use behaviour, access to opioid substitution treatment, and HIV/HCV status in people who inject drugs in Greece by city (Athens and Thessaloniki) [as assessed in their first visit to the programs].

|  | **Overall** | **Athens** | **Thessaloniki** |  |
| --- | --- | --- | --- | --- |
| **Variable** | ***n* = 2433** | ***n* = 1386** | ***n* = 1047** | ***p*-value** |
| Follow-up time (years), median (25<sup>th</sup>–75<sup>th</sup>) | 2.7 (2.2–3.9) | 3.8 (3.0–4.0) | 2.0 (1.6–2.6) | <0.001<sup>1</sup> |
| **A. Socio-demographic and network characteristics** |  |  |  |  |
| Age (years), mean (sd) | 40.1 (8.3) | 39.9 (8.2) | 40.4 (8.4) | 0.127<sup>2</sup> |
| Gender, n (%) |  |  |  | 0.003<sup>3</sup> |
| Female | 375 (15.4) | 240 (17.3) | 135 (12.9) |  |
| Male | 2057 (84.6) | 1146 (82.7) | 911 (87.1) |  |
| Country of origin, n (%) |  |  |  | <0.001<sup>3</sup> |
| Greece | 2167 (89.2) | 1281 (92.6) | 886 (84.6) |  |
| Other | 263 (10.8) | 102 (7.4) | 161 (15.4) |  |
| Highest level of education completed, n (%) |  |  |  | <0.001<sup>3</sup> |
| High school or higher | 902 (37.2) | 596 (43.1) | 306 (29.4) |  |
| Middle/secondary school or below | 1520 (62.8) | 786 (56.9) | 734 (70.6) |  |
| Currently homeless, n (%) |  |  |  | <0.001<sup>3</sup> |
| No | 1965 (80.9) | 1071 (77.4) | 894 (85.6) |  |
| Yes | 463 (19.1) | 312 (22.6) | 151 (14.4) |  |
| Currently health insurance, n (%) |  |  |  | 0.004<sup>3</sup> |
| Yes | 483 (20.0) | 303 (22.0) | 180 (17.4) |  |
| No | 1929 (80.0) | 1072 (78.0) | 857 (82.6) |  |
| Unemployed, n (%) |  |  |  | <0.001<sup>3</sup> |
| Yes | 2130 (87.8) | 1253 (90.7) | 877 (84.0) |  |
| No | 296 (12.2) | 129 (9.3) | 167 (16.0) |  |
| Imprisonment in the past 12 months, n (%) |  |  |  | 0.006<sup>3</sup> |
| No | 2168 (89.3) | 1214 (87.8) | 954 (91.3) |  |
| Yes | 260 (10.7) | 169 (12.2) | 91 (8.7) |  |
| Size of participant's network, n (%) |  |  |  | <0.001<sup>3</sup> |
| ≤10 | 943 (39.1) | 476 (34.7) | 467 (44.9) |  |
| 11–30 | 932 (38.7) | 464 (33.9) | 468 (45.0) |  |
| 30+ | 534 (22.2) | 430 (31.4) | 104 (10.0) |  |
| **B. Injecting drug use behaviour** |  |  |  |  |
| Main substance of use, n (%) |  |  |  | <0.001<sup>3</sup> |
| Heroin/Thai | 1705 (70.3) | 824 (59.6) | 881 (84.5) |  |
| Cocaine/Speedball | 675 (27.8) | 541 (39.1) | 134 (12.9) |  |
| Other | 45 (1.9) | 18 (1.3) | 27 (2.6) |  |
| Injecting drug use in the past 30 days, n (%) |  |  |  | <0.001<sup>3</sup> |
| No | 850 (35.0) | 354 (25.6) | 496 (47.5) |  |
| Yes | 1578 (65.0) | 1029 (74.4) | 549 (52.5) |  |
| Injecting drug use in the past 12 months, n (%) |  |  |  | <0.001<sup>3</sup> |
| No | 344 (14.2) | 85 (6.1) | 259 (24.8) |  |
| Yes | 2084 (85.8) | 1298 (93.9) | 786 (75.2) |  |
| Duration of injecting drug use (years), n (%) |  |  |  | <0.001<sup>3</sup> |
| <18 | 1111 (46.0) | 678 (49.3) | 433 (41.6) |  |
| ≥18 | 1305 (54.0) | 697 (50.7) | 608 (58.4) |  |
| Daily injecting drug use, n (%) |  |  |  | 0.729<sup>3</sup> |
| No | 1759 (72.6) | 1006 (72.9) | 753 (72.3) |  |
| Yes | 663 (27.4) | 374 (27.1) | 289 (27.7) |  |
| Receptive syringe sharing, n (%) |  |  |  | <0.001<sup>3</sup> |
| Never | 1914 (79.2) | 1046 (76.2) | 868 (83.2) |  |
| At least once | 502 (20.8) | 327 (23.8) | 175 (16.8) |  |
| Use drugs divided with a syringe that someone else had already injected with, n (%) |  |  |  | 0.218<sup>3</sup> |
| Never | 2006 (82.8) | 1130 (82.0) | 876 (83.9) |  |
| At least once | 416 (17.2) | 248 (18.0) | 168 (16.1) |  |
| Currently on OST, n (%) |  |  |  | 0.005<sup>3</sup> |
| Yes | 583 (24.0) | 361 (26.1) | 222 (21.3) |  |
| No | 1842 (76.0) | 1020 (73.9) | 822 (78.7) |  |
| anti-HIV, n (%) |  |  |  | <0.001<sup>3</sup> |
| (-) | 2130 (87.6) | 1148 (82.9) | 982 (93.8) |  |
| (+) | 301 (12.4) | 236 (17.1) | 65 (6.2) |  |
| anti-HCV, n (%) |  |  |  | <0.001<sup>3</sup> |
| (-) | 661 (27.2) | 278 (20.1) | 383 (36.6) |  |
| (+) | 1770 (72.8) | 1106 (79.9) | 664 (63.4) |  |
| Coinfection [anti-HIV(+) & anti-HCV(+)], n (%) |  |  |  | <0.001<sup>3</sup> |
| No | 2144 (88.2) | 1153 (83.3) | 991 (94.7) |  |
| Yes | 287 (11.8) | 231 (16.7) | 56 (5.3) |  |
| Chronic HCV, n (%) |  |  |  | <0.001<sup>3</sup> |
| No | 1031 (47.0) | 430 (36.7) | 601 (58.8) |  |
| Yes | 1163 (53.0) | 742 (63.3) | 421 (41.2) |  |
| Chronic HCV with anti-HIV(+), n (%) |  |  |  | <0.001<sup>3</sup> |
| No | 2184 (90.0) | 1177 (85.1) | 1007 (96.5) |  |
| Yes | 243 (10.0) | 206 (14.9) | 37 (3.5) |  |
| First participation based on first national lockdown (15/3/2020) |  |  |  | <0.001<sup>3</sup> |
| Before | 1953 (80.3) | 1386 (100.0) | 567 (54.2) |  |
| After | 480 (19.7) | 0 (0.0) | 480 (45.8) |  |
| Year of entry (first participation) in the study |  |  |  | <0.001<sup>3</sup> |
| 2018 | 1086 (44.6) | 1086 (78.4) | 0 (0.0) |  |
| 2019 | 682 (28.0) | 261 (18.8) | 421 (40.2) |  |
| 2020 | 508 (20.9) | 39 (2.8) | 469 (44.8) |  |
| 2021 | 157 (6.5) | 0 (0.0) | 157 (15.0) |  |

<sup>1</sup>Mann–Whitney U test, <sup>2</sup>Student's t-test, <sup>3</sup>Chi-squared test

**Supplementary Table 2.**
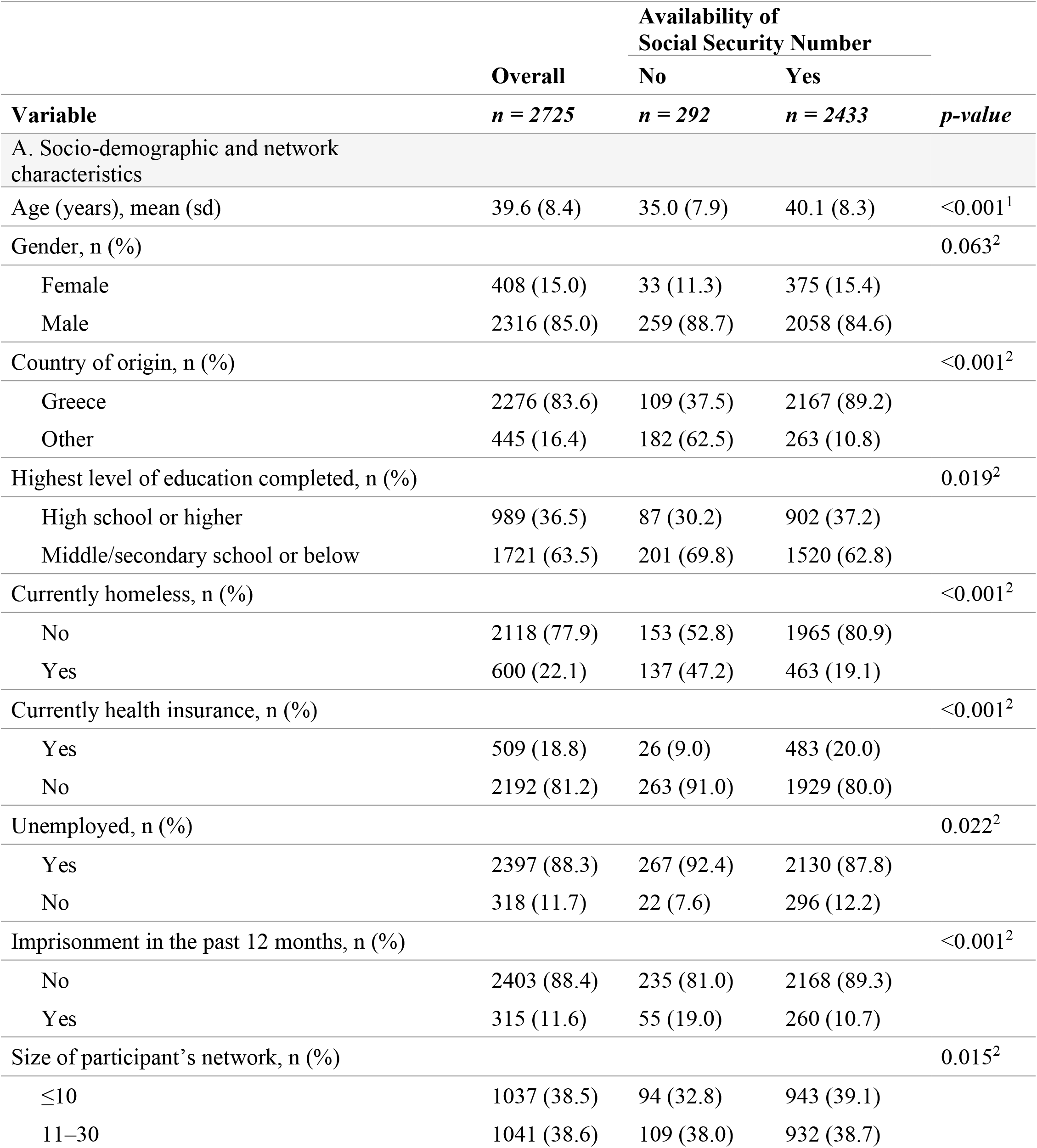
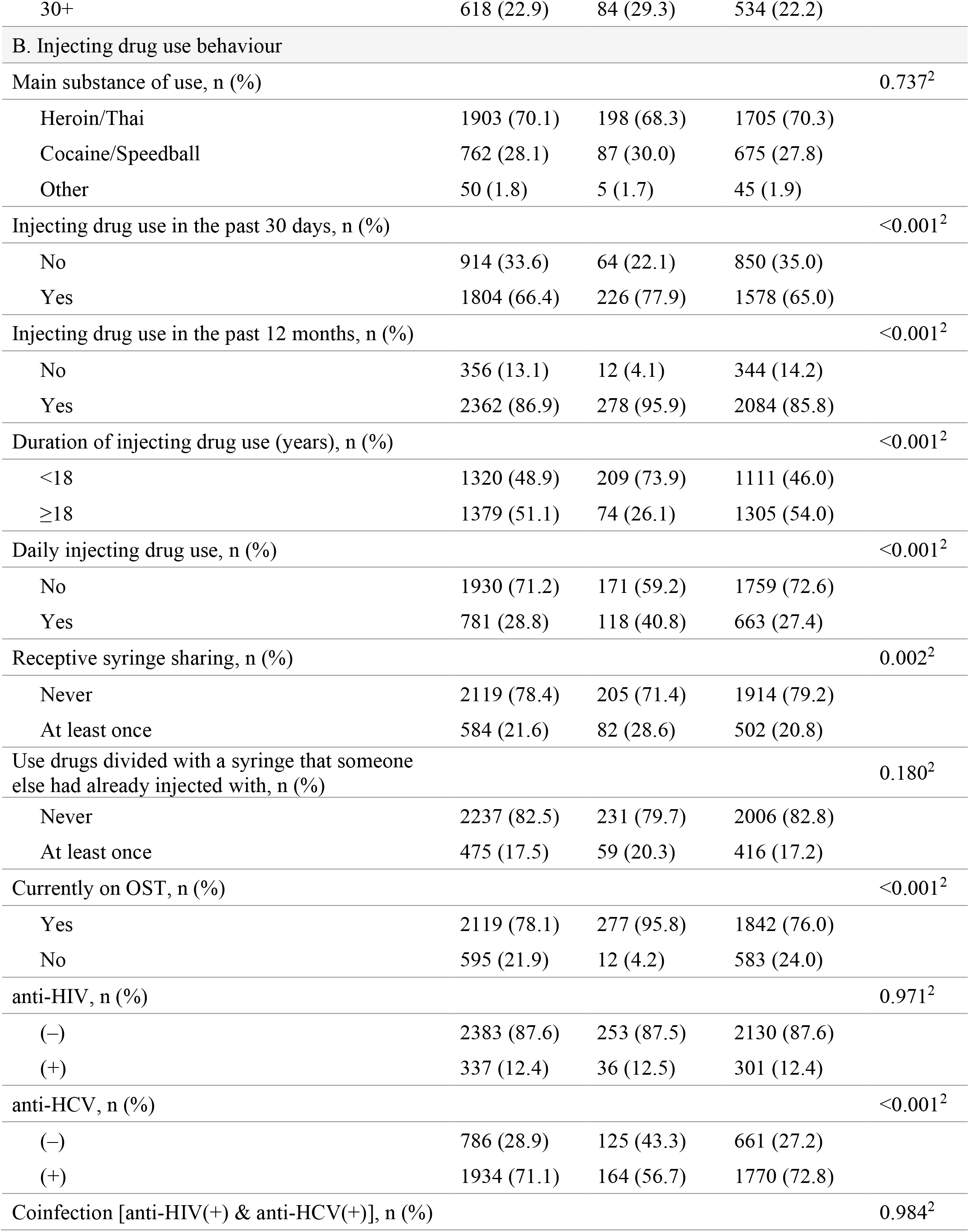
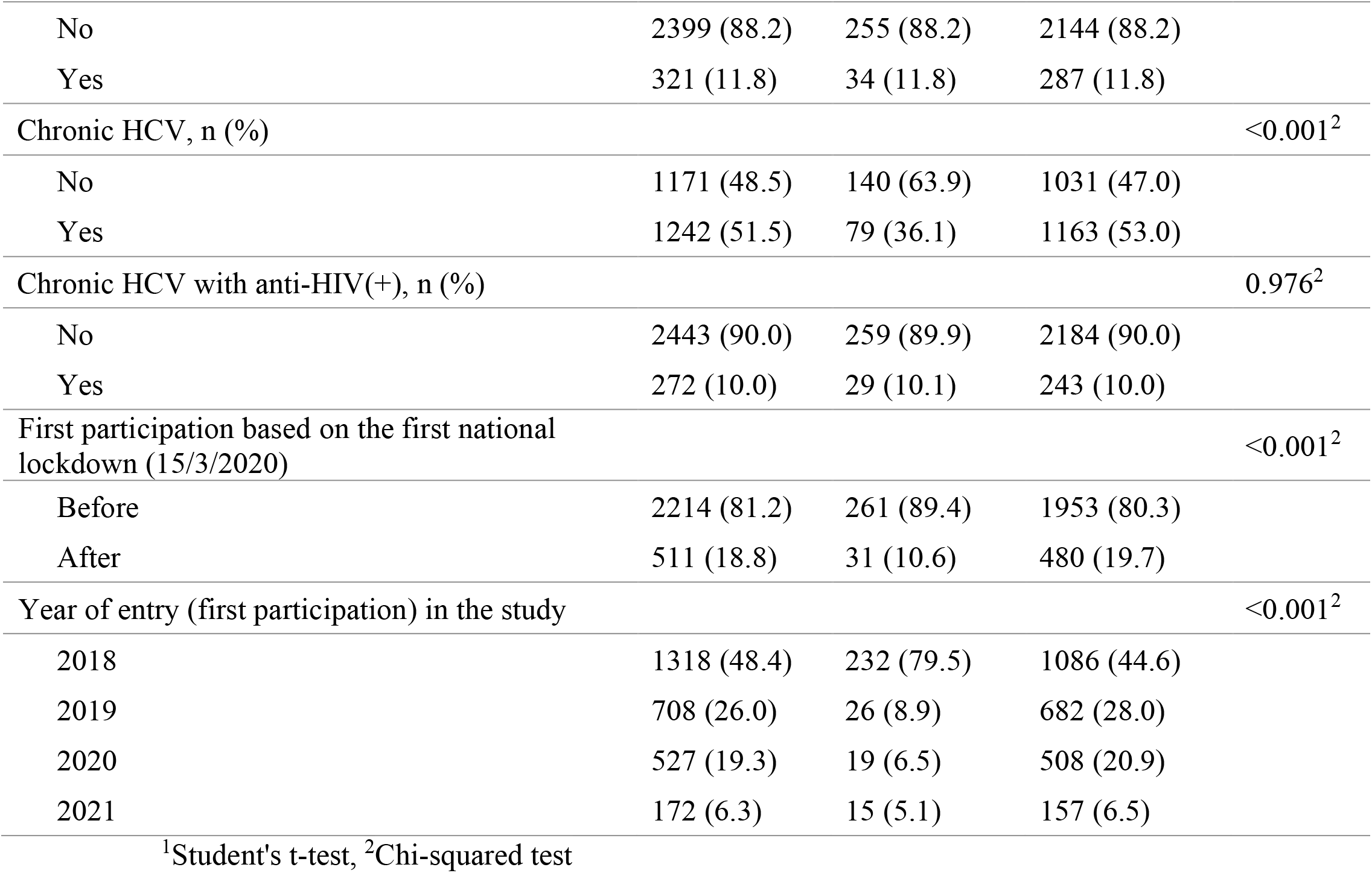
Socio-economic and network characteristics, drug use behaviour, access to opioid substitution treatment, and HIV/HCV status in people who inject drugs by the availability of social security number.

**Supplementary Table 3.** Crude mortality rate (CMR) per 100 person-years by calendar year (overall and by city).

| **All** |  |  | **Athens** |  |  | **Thessaloniki** |  |  |
| --- | --- | --- | --- | --- | --- | --- | --- | --- |
| **Period** | **No.**<br>**Deaths** | **CMR** | **Period** | **No.**<br>**Deaths** | **CMR** | **Period** | **No.**<br>**Deaths** | **CMR** |
| Apr–Dec 2018 | 14 | 2.60 (1.54–4.40) | Apr–Dec 2018 | 14 | 2.60 (1.54–4.40) | - | - | - |
| Jan–Dec 2019 | 28 | 2.29 (1.58–3.32) | Jan–Dec 2019 | 24 | 2.11 (1.41–3.15) | Sep–Dec 2019 | 4 | 4.85 (1.82–12.93) |
| Jan–Dec 2020 | 63 | 3.19 (2.49–4.09) | Jan–Dec 2020 | 40 | 3.01 (2.21–4.11) | Jan–Dec 2020 | 23 | 3.56 (2.36–5.35) |
| Jan–Dec 2021 | 92 | 4.11 (3.35–5.05) | Jan–Dec 2021 | 42 | 3.26 (2.41–4.41) | Jan–Dec 2021 | 50 | 5.27 (4.00–6.96) |
| Jan–Jun 2022 | 46 | 4.69 (3.51–6.26) | Jan–Jun 2022 | 27 | 4.87 (3.34–7.10) | Jan– Jun 2022 | 19 | 4.46 (2.84–6.99) |
| Overall | 243 | 3.50 (3.08–3.97) | Overall | 147 | 3.03 (2.58–3.57) | Overall | 96 | 4.56 (3.74–5.57) |

**Supplementary Figure 1.**
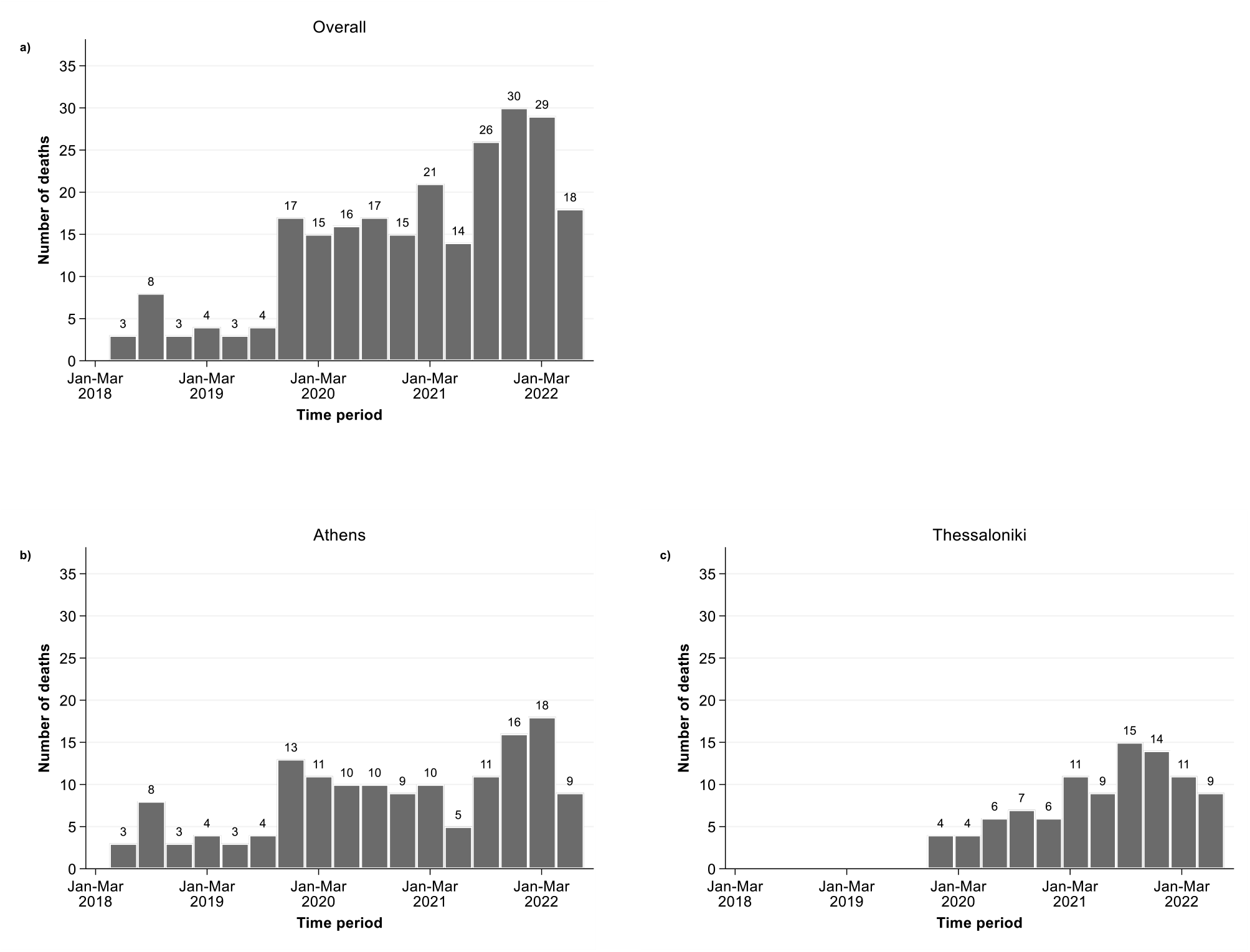
Distribution of deaths per quarter a) Overall, b) Athens, and c) Thessaloniki

